# Development and Validation of MyProstateScore 2.0 to Detect Clinically Significant Prostate Cancer

**DOI:** 10.1101/2023.04.11.23288418

**Authors:** Jeffrey J. Tosoian, Yuping Zhang, Lanbo Xiao, Cassie Xie, Nathan L. Samora, Yashar S. Niknafs, Zoey Chopra, Javed Siddiqui, Heng Zheng, Grace Herron, Neil Vaishampayan, Kumaran Arivoli, Bruce J. Trock, Ashley E. Ross, Todd M. Morgan, Ganesh S. Palapattu, Simpa S. Salami, Lakshmi P. Kunju, Yingye Zheng, John T. Wei, Arul M. Chinnaiyan

## Abstract

**Background:** The benefits of prostate cancer screening with serum prostate-specific antigen (PSA) have been largely offset by the high rate of negative prostate biopsies and overdiagnosis of indolent cancers. These outcomes result from the limited diagnostic accuracy of PSA for clinically significant prostate cancer and are considered a major obstacle to realizing the benefits of population-wide screening for prostate cancer.

**Methods:** We analyzed RNAseq data from a prostate cancer compendium to identify novel transcripts associated with cancer and high-grade cancer. Predefined nomination criteria were applied to 58,724 gene targets, yielding 54 differentially expressed transcripts. We designed a custom multiplex qPCR panel for non-invasive detection of candidate transcripts in urine. The panel was applied to a development cohort of men with elevated PSA (3 to 10 ng per milliliter) that underwent prospective, standardized urine collection and prostate biopsy at the University of Michigan. Elastic net modeling was used to derive the optimal model for clinically significant (grade group 2 or higher) prostate cancer, the 18-transcript MyProstateScore 2.0 (MPS2) test. The calibrated, locked MPS2 model was assessed in a blinded, external National Cancer Institute (NCI) – Early Detection Research Network (EDRN) validation cohort and compared to serum PSA, the Prostate Cancer Prevention Trial risk calculator (PCPTrc), and the MyProstateScore (MPS) test. The original MPS assay measures urinary expression of two cancer-associated markers (PCA3, TMPRSS2:ERG) and is endorsed by National Comprehensive Cancer Network guidelines for consideration prior to biopsy in the study population.

**Results:** We performed multiplex urinary testing of 1,623 clinical specimens in total, representing the largest such cohort to our knowledge. The prospective NCI-EDRN validation population included 743 men undergoing per-protocol urine collection and prostate biopsy. The median age was 62 years, median PSA was 5.6 ng per milliliter, and 151 men (20%) had clinically significant prostate cancer on biopsy. The area under the receiver-operating characteristic curve (AUC) for clinically significant prostate cancer was 0.597 (95% Confidence Interval [CI], 0.547 to 0.646) for PSA, 0.659 (95% CI, 0.611 to 0.707) for the PCPTrc, and 0.737 (95% CI, 0.694 to 0.780) for MPS, as compared to 0.818 (95% CI, 0.781 to 0.855) for the optimal MPS2 model (MPS2+). Under a clinically applicable testing approach providing 95% sensitivity for clinically significant cancer, the specificity (equivalent to the percentage of unnecessary biopsies avoided after pre-biopsy testing) was 11% for PSA, 20% for the PCPTrc, and 23% for MPS, as compared to 41% for MPS2+. In all sub-populations, MPS2 testing provided negative predictive value (NPV) of 95% to 99% for clinically significant cancer.

**Conclusions:** In a large, external validation population referred for prostate biopsy, the novel MPS2 assay provided exceptional sensitivity and NPV to rule out clinically significant prostate cancer. These data support the use of MPS2 as a highly accurate secondary test to reduce the harms associated with PSA screening and preserve its long-term benefits.

## INTRODUCTION

Prostate cancer remains the most commonly diagnosed malignancy and a leading cause of cancer death in the developed world.^1^ The European Randomized Study of Screening for Prostate Cancer (ERSPC) and randomized GOTEBORG-1 trial demonstrated significant reductions in prostate cancer mortality among men participating in prostate-specific antigen (PSA)-based screening followed by transrectal ultrasound (TRUS)-guided prostate biopsy.^2–5^ At the same time, these data revealed that PSA screening led to a high rate of invasive biopsies in men without prostate cancer and overdiagnosis of low-grade, indolent cancers (grade group 1).

These negative outcomes of screening are a direct result of the low specificity of PSA for prostate cancer. A product of prostatic epithelial cells, the PSA protein is expressed by both normal and neoplastic prostate tissue. As a result, PSA levels are elevated for a variety of non-neoplastic causes. Under the traditional clinical pathway, in which men with a PSA level of 3 ng per milliliter or greater undergo systematic prostate biopsy, an estimated 84% of biopsies performed are found to be negative or overdiagnose low-grade, indolent cancer.^5^ The frequency of unnecessary biopsies performed and low-grade cancers overdiagnosed resulting from use of PSA as an isolated screening test is considered a major obstacle to acceptance of population-based screening for prostate cancer, despite its potential benefits.^6–9^ In response to this, contemporary clinical guidelines recommend evaluation prior to prostate biopsy with multiparametric magnetic resonance imaging (mpMRI), if available, and consideration of biomarker testing.^10^

However, the use of mpMRI as a first-line test after PSA screening has significant limitations. High-quality data from expert centers demonstrate a modest 91% pooled negative predictive value (NPV) for clinically significant cancer (grade group 2 or higher).^11^ Furthermore, mpMRI interpretation is highly reader-dependent, with NPV ranging as low as 63% at some academic centers and as low as 40% among individual radiologists.^11, 12^ Thus, even following a negative mpMRI, the persistent risk of clinically significant cancer merits proceeding to biopsy in a substantial proportion of the population. Moreover, there are several practical reasons that mpMRI is not well-suited as a first-line test after PSA screening, including its high cost, time and resource burden, and limited availability to patients.^13, 14^

An alternative solution is first-line use of objective, non-invasive biomarker tests obtainable in the course of routine clinical care. Several blood- and urine-based assays incorporating cancer-specific biomarkers have consistently outperformed PSA-based tools in detection of clinically significant cancer. One example is the MyProstateScore test (MPS), which measures expression of prostate cancer antigen 3 (PCA3) and the TMPRSS2:ERG (T2:ERG) gene fusion in clinical urine specimens.^15^ The multivariable MPS model has outperformed PSA-based tools in detecting clinically significant prostate cancer across multiple validation studies^16, 17^ and is currently proposed by the National Comprehensive Cancer Network (NCCN) for consideration prior to biopsy in men with elevated PSA.^10^

Acknowledging the indolent nature of low-grade prostate cancer, contemporary guidelines emphasize a narrowed clinical focus on detection of higher-grade, clinically significant cancers.^10, 18, 19^ While cancer-specific biomarkers introduced in the last two decades have improved upon PSA, currently available assays have not evolved to reflect this monumental shift in the clinical understanding of prostate cancer. We therefore sought to identify novel biomarkers specifically overexpressed by high-grade, potentially lethal prostate cancers. Leveraging our own transcriptomic studies of prostate tumors^20, 21^ and those of the public domain,^22^ we hypothesized that augmenting the two-marker MPS assay with biomarkers specifically linked to high-grade prostate cancer would improve upon a past generation of markers expressed by indolent and aggressive cancers alike. Translating our findings to a practical testing platform for clinical adoption, we developed a novel non-invasive assay for clinically significant prostate cancer and externally validated its performance relative to other currently available testing options.

## METHODS

### RNA SEQ-BASED TRANSCRIPT DISCOVERY

Having characterized the diagnostic accuracy of the two-marker MPS assay,^16^ we sought improved detection of clinically significant prostate cancer through the addition of novel transcripts specifically over- or under-expressed by high-grade cancers. Our discovery analysis included tissue-based RNA sequencing (RNAseq) data available through The Cancer Genome Atlas (TCGA) consortium,^22^ the Genotype-Tissue Expression (GTEx) portal,^23^ and the University of Michigan (UM).^20, 21, 24^ The discovery set (N=775) included 220 normal (benign) prostates, 71 low-grade cancers (grade group 1), and 484 higher-grade, clinically significant cancers (grade group 2 or higher).

The analysis evaluated 58,724 gene targets (**Supplementary Appendix Table S1**). To identify genes with high potential for subsequent non-invasive detection, we prioritized transcripts with high absolute expression in cancer and high-grade cancer in addition to differential expression across benign, low-grade, and high-grade cancers. Gene quantification was performed using the GENCODE v29 transcriptome.^25^ RNAseq data were processed using the Kallisto gene quantification tool (v0.44.0).^26^ Analyses were performed in R, using edgeR^27^ for normalization and limma-voom for differential expression testing.^28^ Clustering statistics were performed using log-normalized transcripts per million (TPM) values to classify genes as high-expressing or low-expressing for benign, low-grade cancer, and high-grade cancer. Nomination criteria for differentially expressed high-grade genes were defined *a priori* and included: i) gene length >500 base pairs, ii) differential expression adjusted p-value <0.1, iii) log_2_ mean fold-change >0, iv) >85% of high-expressing samples in high-grade cancers via clustering statistics, v) log_2_ fold-change of high-expressing sample mean to low-expressing sample mean >0, vi) 95^th^percentile normal expression <50 TPM, and vii) 95^th^ percentile high-grade expression >30 TPM. Additional nomination criteria are detailed in the **Supplementary Appendix Table S2**.

### DEVELOPMENT COHORT

Urinary specimens have been prospectively collected prior to prostate biopsy at the UM Prostate SPORE under an IRB-approved, National Cancer Institute-Early Detection Research Network (NCI-EDRN) standardized protocol since 2008. First-catch urine is obtained following digital rectal examination (DRE), mixed with RNA stabilization buffer, and frozen to -70°C as described.^29^ The development cohort was comprised of patients presenting to UM for prostate biopsy due to elevated PSA and/or suspicious DRE from 2008 through 2020. In accordance with clinical guidelines for intended use of prostate cancer biomarkers,^10^ we excluded patients with a previous diagnosis of prostate cancer, serum PSA level less than 3 ng per milliliter or greater than 10 ng per milliliter, or a history of pre-biopsy prostate mpMRI. All patients underwent TRUS-guided systematic biopsy of 12 or more cores, and biopsies were graded in accordance with the International Society of Urological Pathology (ISUP) Consensus Conference.^30, 31^

### VALIDATION COHORT

The validation cohort consisted of patients participating in the prospective NCI-EDRN PCA3 Evaluation Trial. This trial enrolled a consecutive series of patients presenting for prostate biopsy at one of eleven academic centers, primarily due to elevated or increasing PSA or abnormal DRE, as previously described and detailed in the **Supplementary Appendix Table S3**.^29^ Additional eligibility criteria for the current study included availability of sufficient urine volume and clinical data for analysis. Specimen collection was performed per the identical NCI-EDRN protocol employed at UM, and all patients underwent TRUS-guided systematic biopsy, of which 736 (99%) included 12 or more cores. Pathologic interpretation was performed by genitourinary pathologists at each respective academic center in accordance with the ISUP Consensus Conference.^30, 31^ A randomized 10% of sample specimens were independently re-reviewed by central pathology. All participants provided informed consent.

### LABORATORY PROCEDURES

#### Multiplex Quantitative-PCR OpenArray™ Profiling

OpenArray™ technology (Thermo Fisher Scientific, Waltham, MA, USA) is a high-throughput real-time quantitative polymerase chain reaction (qPCR) method that allows for rapid screening of multiple TaqMan™ assays across samples. This real-time method uses an array of 3,072 through-holes run on the QuantStudio 12K Flex Real-Time PCR System with an OpenArray™ block. RNA isolation for the 54-gene OpenArray™ panel was performed using the MagMAX™ mirVana™ Total RNA Isolation Kit. Briefly, 500 microliters of a 1 to 1 mixture of urine and Hologic transport media were mixed 1 to 1 with Lysis Binding Mix. Binding Beads Mix was then added to enrich nucleic acids, followed by TURBO DNase digestion and RNA elution. For high-throughput RNA extraction, urine samples were processed through the semi-automatic KingFisher Flex System (Thermo Fisher Scientific).

After RNA extraction, 16 microliters of RNA were used to synthesize cDNA using SuperScript™ IV VILO™ Master Mix (Thermo Fisher Scientific), followed by pre-amplification with TaqMan™ PreAmp Master Mix (Thermo Fisher Scientific). For each sample, 2.5 microliters of pre-amplified cDNA and 2.5 microliters of 2× TaqMan OpenArray™ Master Mix were mixed and loaded into 384-well plates per manufacturer instructions. The QuantStudio 12K Flex OpenArray™ AccuFill System transferred the previously generated mix to the TaqMan OpenArray™ plate. Amplification was performed using the QuantStudio 12K Flex Real Time PCR System, and the delta-delta cycle threshold method was used for analysis with the QuantStudio™ 12K Flex Software. All samples were run in triplicate.

### BIOINFORMATIC AND STATISTICAL ANALYSIS

#### Data Pre-Processing

The measure of gene expression was the cycle threshold (Ct), defined as the number of amplification cycles required for sample fluorescence to exceed background level. Ct values are inversely related to the quantity of nucleic acid in a sample, with lower Ct values reflecting increased expression of the target gene. Expression data were pre-processed to account for outlier and undetermined replicates, and the mean Ct was determined based on valid replicates. The mean Ct of each target marker was normalized to the housekeeping gene *KLK3* using the formula -[*Ct_target_–Ct_KLK3_*], and the normalized mean Ct was used for model building.

#### MyProstateScore Calculation

MPS results in the development and validation cohorts were calculated by entering normalized Ct values for PCA3 and T2:ERG from OpenArray™ quantification into the original, locked MPS model previously described.^15^

#### Model Development

The primary outcome was clinically significant prostate cancer, defined as grade group 2 or higher cancer. Normalized mean Ct values of gene targets were assessed with pertinent clinical variables to develop a novel model expanding on the two-marker MPS test, the MyProstateScore 2.0 (MPS2) test. Clinical factors consistently associated with clinically significant prostate cancer and available at no cost (age, race, family history of prostate cancer, abnormal DRE, and prior negative biopsy) were locked into the model *a priori*.^32^ Notably, inclusion of prostate volume in clinical models has improved discrimination for clinically significant prostate cancer,^33, 34^ however prostate volume is not readily available in all patients undergoing clinical risk assessment. As such, to provide each patient with optimal risk quantification using available clinical data, we developed a parallel model incorporating prostate volume, the MyProstateScore 2.0 plus (MPS2+) model, for its use when prostate volume is clinically available.

Based on analysis of optimal feature size and technical features of the OpenArray™ platform, the final model could include up to 18 candidate markers improving discrimination for clinically significant prostate cancer. Input variables were assessed for collinearity using a stepwise approach. Specifically, variance inflation factor (VIF) was calculated for all variables, and the variable with the highest VIF was removed. This was repeated until no variables remained with VIF >5, resulting in the exclusion of nine collinear markers. We evaluated three model-building strategies: i) logistic regression with stepwise feature selection, ii) logistic regression with recursive feature elimination (RFE), and iii) regularized logistic regression with elastic net. Model performance was quantified as the area under the receiver-operating characteristic (ROC) curve (AUC) on repeat cross-validation (10-fold cross-validation repeated three times) with upsampling of the minor class (clinically significant prostate cancer) to create balanced classes for development. Across all analyses, elastic net modeling demonstrated the highest median AUC and was thus used for model development. Acknowledging the potential for misclassification of study outcome based on biopsy undersampling, we performed a sensitivity analysis of model development in which subjects with additional pathologic assessment (e.g., radical prostatectomy pathology) were classified by the highest-grade cancer detected (**Supplementary Appendix Table S4**). Model evaluation was performed using the *train* function with method set to *glmStepAIC*, *rfe*, or *glmnet* in the R package *caret*.^35^

#### Model Calibration

Calibration is necessary when there is distributional shift in the outcome between development and validation populations, and its importance for clinical modeling has been described in detail.^36^ As described, the minor outcome class (clinically significant prostate cancer) was upsampled to yield balanced classes in development, while the validation cohort was a consecutive series of patients indicated for prostate biopsy, consistent with the clinical population appropriate for biomarker testing. Two calibration methods were applied to a re-sampled development set with matched outcome prevalence: i) recalibration in the large, which includes re-estimation of the model intercept, and ii) logistic recalibration, which includes re-estimation of model intercept and slope.^37^ The latter method provided superior performance and was used for calibration as previously described.^38^ Pre- and post-calibration curves were generated using the *calibration* function from the R package *caret*.

#### Blinded, External Model Validation

To evaluate the robustness of MPS2 and MPS2+, the calibrated, locked models were assessed in the NCI-EDRN external validation cohort. De-identified specimens were shipped to the University of Michigan for OpenArray™ profiling. Laboratory procedures and data pre-processing were conducted per the identical protocol used in development. Normalized mean Ct values of the target markers and the locked MPS2 coefficients were provided to the EDRN Data Management and Coordinating Center for external assessment. Notably, NCI-EDRN cohort data were accessible by only two investigators (CX, Y. Zheng), who performed the validation analysis.

In the external validation cohort, the overall discriminative accuracy (AUC) of PSA, the PSA-based Prostate Cancer Prevention Trial risk calculator (PCPTrc, which includes serum PSA, age, race, family history of prostate cancer, DRE findings, and history of prior negative biopsy),^32^ the original MPS test,^15^ and the locked MPS2 models were plotted. Threshold-based analyses were rooted in the clinical importance of ruling out clinically significant cancer with high sensitivity and NPV,^39^ i.e. minimizing false negative tests, thereby enabling patients and clinicians to confidently forego additional testing. Thus, our primary analysis considered threshold values providing 95% sensitivity, and the associated specificity, NPV, and positive predictive value (PPV) were calculated for each test. To illustrate the clinical impact of each test, we calculated the percentage of unnecessary biopsies (biopsies negative for cancer or overdiagnosing grade group 1 cancer) avoided based on use of each test to select for biopsy.

Decision curve analysis (DCA) was used to quantify the net benefit provided by each test on the decision to undergo prostate biopsy as compared to: i) biopsying all patients and ii) biopsying no patients. Considering clinically significant cancer risk exceeding 20% justifies performing biopsy in most patients and risk less than 5% justifies foregoing biopsy in most patients,^40^ we considered threshold probabilities spanning this clinically applicable range of risk. DCA was performed using the *dca* function in the R package *dcurves*. All statistical analyses were performed using R version 4.1.

## RESULTS

### NOMINATION OF TRANSCRIPTS ASSOCIATED WITH HIGH-GRADE PROSTATE CANCER

The first aim of this study was to identify biologic markers differentially expressed by prostate cancer and higher-grade, clinically significant prostate cancer. We performed differential expression analysis of 58,724 genetic targets in RNAseq data derived from a prostate cancer transcriptome compendia spanning UM and the public domain (**Supplementary Appendix Table S1**). Application of predefined nomination criteria for cancer and high-grade cancer yielded 72 genes (**Figure 1A**). Exclusion of genes with significant collinearity and those for which reliable PCR primers could not be feasibly designed yielded 44 candidate genes (**Supplementary Appendix Figures S1-S3**). The candidate pool was supplemented with ten markers with a previously reported association with cancer or suitability for normalization. This yielded a final development panel of 54 unique genetic targets, including 17 high-grade cancer-associated genes, 27 cancer-associated genes, and ten curated genes (**Supplementary Appendix Table S5**).

**Figure 1.**
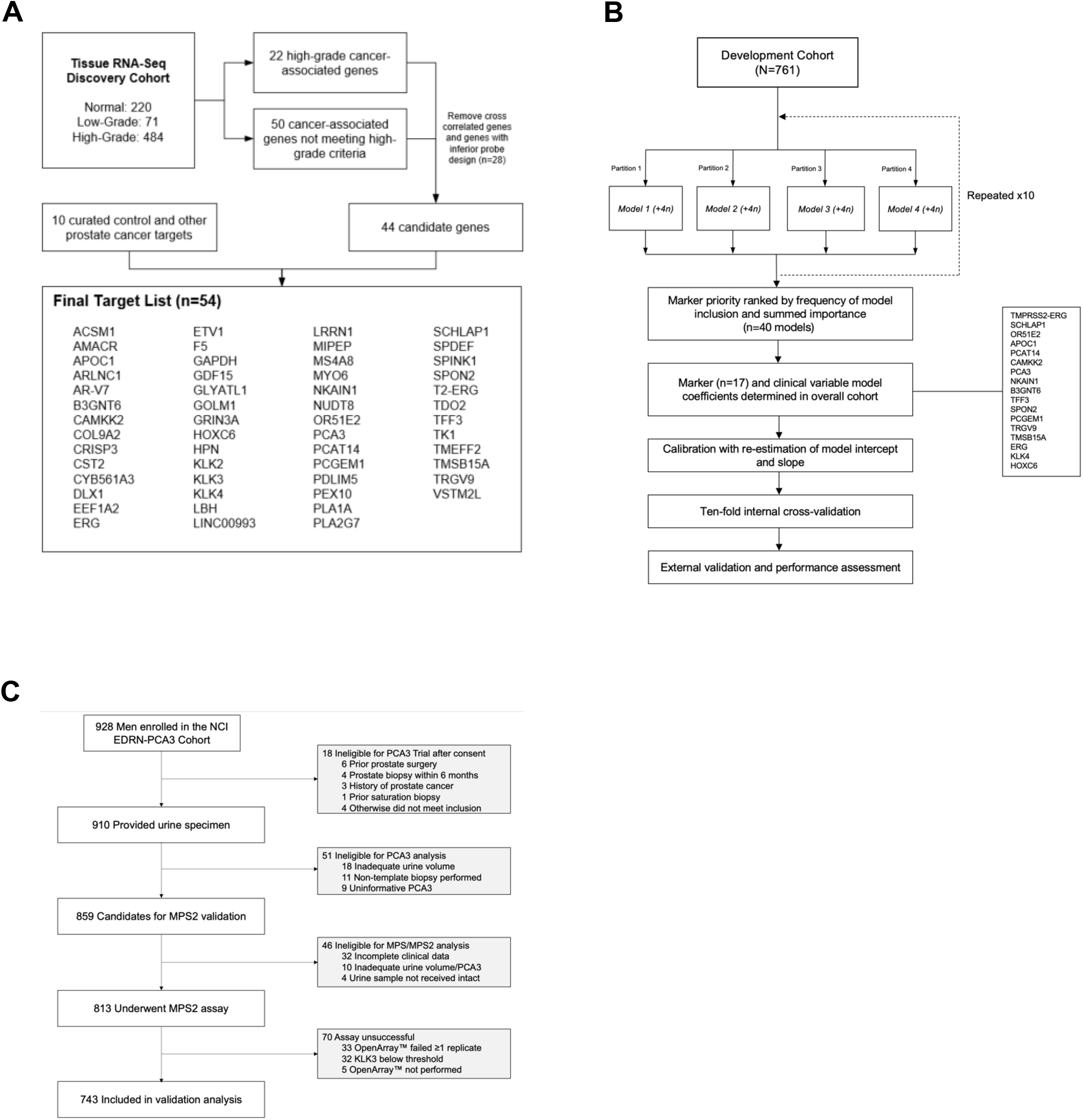
Discovery, Development, and Validation of the MyProstateScore 2.0 (MPS2) Urinary Test for Detection of Clinically Significant Prostate Cancer. Shown is the nomination and selection pathway for RNA transcript inclusion in the multiplex qPCR candidate panel (Panel A). RNAseq data from TCGA, GTEx, and UM prostate tumor studies were assessed. Forty-four candidate transcripts were nominated by tissue RNAseq analysis and combined with 10 curated control and prostate cancer-associated targets (total of 54 gene targets). Shown is the model development pipeline applied to urine multiplex qPCR data from the UM development cohort (Panel B). Model building employed an ensemble approach, in which the importance of candidate markers to model accuracy across 40 resamplings was determined by elastic net analysis. The 17 biomarkers providing optimal discriminative accuracy for clinically significant prostate cancer were added to standard clinical variables and the normalization gene *KLK3* in the MPS2 and MPS2+ (plus prostate volume) models. Models were calibrated and internally cross-validated prior to external validation. Shown is the external validation cohort comprised of men undergoing prostate biopsy in the National Cancer Institute – Early Detection Research Network (NCI-EDRN) PCA3 Trial (Panel C). Of 859 men participating in the PCA3 trial, 46 (5.4%) were ineligible for the current analysis due to inadequate urine volume or unavailable clinical data. Of 813 eligible participants, the MPS2 assay was successfully performed in 743 (91%), yielding the final external validation population.

### DEVELOPMENT OF THE MYPROSTATESCORE 2.0 (MPS2) MODEL

To develop an optimal multiplex model for clinically significant cancer, we turned our attention to the clinical population indicated for diagnostic biomarker testing (i.e., men with serum PSA level greater than 3 ng per milliliter). The development cohort included 815 eligible participants with adequate urine volume for multiplex qPCR (**Supplementary Appendix Figure S4**). Among these, the assay yielded valid expression data in 761 men (93%), of which 293 (39%) had clinically significant prostate cancer on biopsy. Median age (interquartile range, IQR) in the development cohort was 63 years (58 to 68), median PSA was 5.6 ng per milliliter (4.6 to 7.2), and 163 men (21%) had a history of previous negative biopsy (**Table 1**).

**Table 1.** Characteristics of the development and external validation populations overall and stratified by detection of clinically significant prostate cancer.*.

| Characteristic | Development Cohort |  |  | External Validation Cohort |  |  |
| --- | --- | --- | --- | --- | --- | --- |
|  | Total<br>(N=761) | Negative or GG1<br>(N=468) | GG 2 to 5<br>(N=293) | Total<br>(N=743) | Negative or GG1<br>(N=592) | GG 2 to 5<br>(N=151) |
| Median age (IQR) – years | 63 (58-68) | 63 (57-68) | 64 (58-69) | 62 (57-68) | 62 (57-68) | 64 (59-70) |
| African-American – no. (%) | 33 (4.3%) | 16 (3.4%) | 17 (5.8%) | 95 (13%) | 70 (12%) | 25 (17%) |
| Positive family history – no. (%) | 206 (27%) | 116 (25%) | 90 (31%) | 212 (29%) | 164 (28%) | 48 (32%) |
| Previous negative biopsy – no. (%) | 163 (21%) | 127 (27%) | 36 (12%) | 247 (33%) | 229 (39%) | 18 (12%) |
| Abnormal DRE – no. (%) | 104 (14%) | 38 (8.1%) | 66 (23%) | 139 (19%) | 88 (15%) | 51 (34%) |
| Median prostate volume † – ml | 48 (36-66) | 53 (40-72) | 41 (30-54) | 43 (32-60) | 46 (34-65) | 36 (28-47) |
| Median PSA (IQR) – ng/ml | 5.6 (4.6-7.2) | 5.6 (4.6–6.9) | 5.6 (4.7-7.5) | 5.6 (4.1-8.0) | 5.4 (4.0–7.7) | 6.2 (4.7–8.9) |
| Median PSA density ‡ (IQR) – ng/ml<sup>2</sup> | 0.12 (0.08-0.16) | 0.10 (0.07-0.14) | 0.15 (0.10-0.20) | 0.12 (0.08-0.19) | 0.11 (0.08–0.17) | 0.17 (0.12-0.31) |
| Median MPS2 § (IQR) | 0.16 (0.05-0.39) | 0.07 (0.03-0.19) | 0.40 (0.20-0.61) | 0.13 (0.05–0.37) | 0.10 (0.04-0.24) | 0.44 (0.23–0.69) |
| Median MPS2+ § (IQR) | 0.14 (0.05-0.42) | 0.07 (0.03-0.17) | 0.44 (0.22-0.68) | 0.15 (0.05–0.43) | 0.11 (0.04–0.30) | 0.54 (0.27-0.79) |
| Biopsy GG ≥2 – no. (%) | 293 (39%) | 0 | 293 (100%) | 151 (20%) | 0 | 151 (100%) |

\* GG denotes grade group, DRE digital rectal examination, PSA prostate-specific antigen, MPS2 MyProstateScore 2.0, MPS2+ MyProstateScore2.0+.
† Measured by transrectal ultrasound (TRUS).
‡ PSA density equals serum PSA divided by prostate volume.
§ MPS2 and MPS2+ values are reported on a continuous scale as the likelihood of detecting clinically significant prostate cancer on biopsy.

To identify a robust panel of genes for the clinical MPS2 models, we employed an ensemble approach, which integrates information from elastic net models developed in multiple resamplings (**Figure 1B**). The development set was randomly divided into four partitions, and the model deriving maximal AUC was identified for each partition. This approach was repeated ten times with different random seeds, yielding 40 elastic net models in total. The frequency with which markers were selected and their importance to respective models, as determined by elastic net analysis, was calculated. In addition to standard clinical variables (age, race, family history of prostate cancer, DRE findings, and history of prior negative biopsy), the final MPS2 model included 17 markers of clinically significant prostate cancer and the reference gene *KLK3* (**Supplementary Appendix Table S6**). Prostate volume was included to yield the MPS2+ model in parallel. After logistic re-calibration, post-calibration curves confirmed the final models were well-calibrated for risk of clinically significant cancer (**Supplementary Appendix Figure S5A-5B**).

### INTERNAL ASSESSMENT AND VALIDATION OF THE MPS2 MODELS

In the development cohort, median MPS2 values were significantly higher in men with clinically significant prostate cancer (0.40, IQR 0.20 to 0.61) than those with negative biopsies or low-grade cancer (0.07, IQR 0.03 to 0.19, p<0.001) (**Table 1**, **Figure 2A**). Similarly, median MPS2+ was 0.44 (IQR 0.22 to 0.68) in men with clinically significant cancer as compared to 0.07 (IQR 0.03 to 0.17, p<0.001) in those with negative biopsies or low-grade cancer. The ROC curves of PSA, the PCPTrc, and MPS were calculated in the development cohort for comparison (**Figure 2B**). For an unbiased assessment of model building in the development cohort, ROC curves from each cross-validation fold and the mean ROC were plotted for MPS2 and MPS2+ (**Figure 2C-D**). Overall diagnostic accuracy (AUC) values for clinically significant prostate cancer were 0.539 for PSA, 0.609 for the PCPTrc, and 0.724 for MPS, as compared to 0.802 for MPS2 and 0.821 for MPS2+.

**Figure 2.**
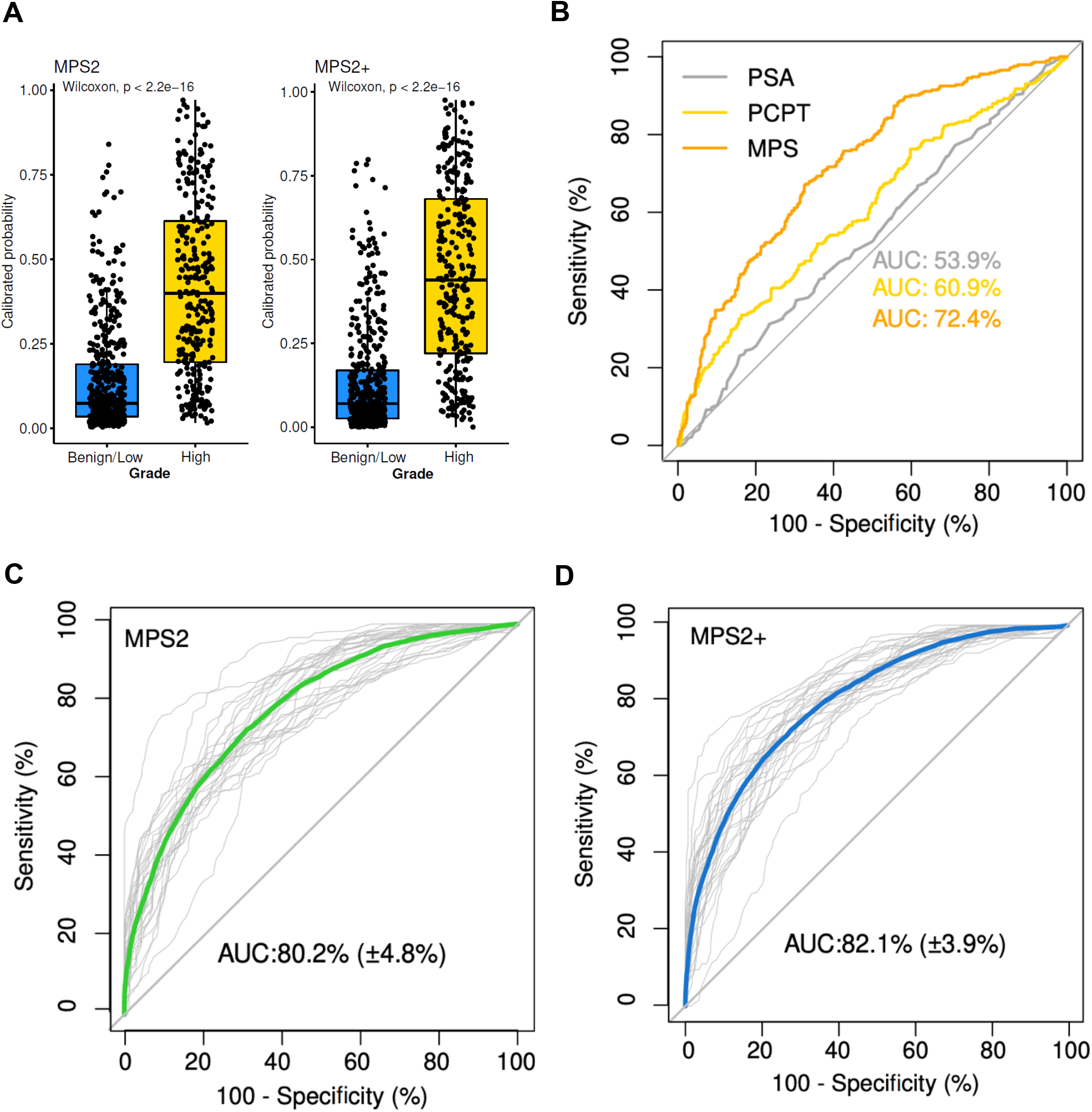
Performance of the Cross-Validated MPS2 Models for Clinically Significant Prostate Cancer in the Development Cohort. Shown are box and dot plots illustrating the distribution of MPS2 and MPS2+ values in men without (blue) and with (yellow) clinically significant prostate cancer in the development cohort (Panel A). Median MPS2 and MPS2+ were significantly higher in men with clinically significant cancer compared to men with no cancer or grade group 1 cancer (p<0.001 for both comparisons). Shown are the receiver-operating characteristic (ROC) curves and the corresponding areas under the curve (AUC) for PSA (gray), the PCPT risk calculator (yellow), and MPS (orange) (Panel B), and the cross-validated MPS2 (Panel C) and MPS2+ (Panel D) tests. Panels C and D illustrate ROC curves of individual cross-validation folds (thin gray lines) and the mean ROC of all cross-validation folds for MPS2 (green) and MPS2+ (blue).

### EXTERNAL VALIDATION OF THE MPS2 MODELS

Of 928 men consented for protocolized urine collection and prostate biopsy in the NCI-EDRN PCA3 Evaluation trial,^29^ 859 (93%) were eligible for the primary (i.e., PCA3) analysis. Among them, 46 (5.4%) were ineligible for the current study due to inadequate urine volume or unavailable clinical data (**Figure 1C**). The remaining 813 subjects underwent OpenArray™ testing, which yielded valid results in 743 (91%), making up the final validation cohort. The median participant age was 62 years (IQR 57 to 68), median PSA was 5.6 ng per milliliter (IQR 4.1 to 8.0), and 247 men (33%) had previously undergone a negative biopsy. On study biopsy, 151 men (20%) were found to have clinically significant prostate cancer (**Table 1**).

The median MPS2 was 0.10 (IQR 0.04 to 0.24) in men with no cancer or grade group 1 cancer on biopsy as compared to 0.44 (IQR 0.23 to 0.69) in men with clinically significant cancer (**Figure 3A**, p<0.001). Similarly, MPS2+ was significantly higher in men with clinically significant cancer than men with no cancer or grade group 1 cancer (median 0.54 vs. 0.11, p<0.001).

**Figure 3.**
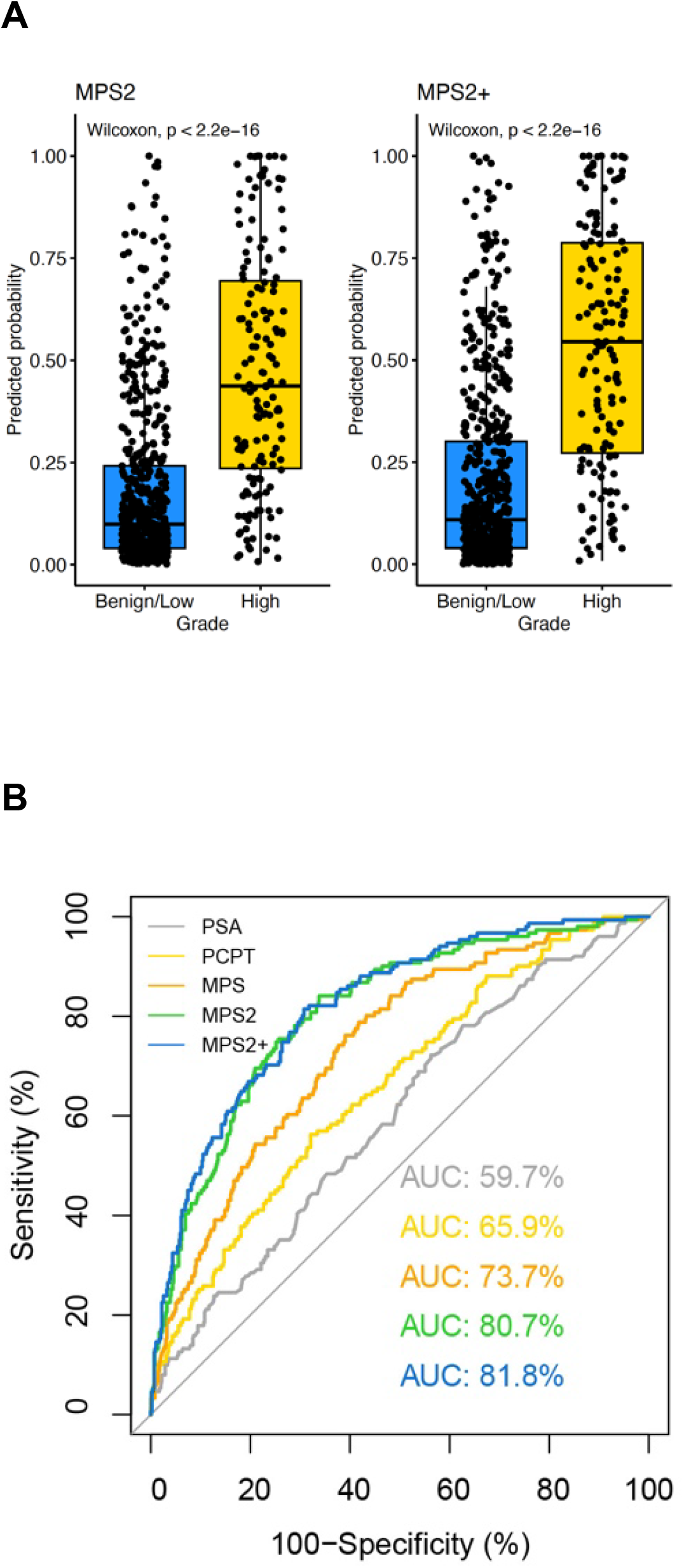
Performance of the Locked MPS2 Models in the External Validation Cohort. Shown are box and dot plots illustrating the distribution of MPS2 and MPS2+ values in men without (blue) and with (yellow) clinically significant prostate cancer in the external validation cohort (Panel A), and receiver-operating characteristic curves and areas under the curve (AUC) for PSA (gray), the PCPT risk calculator (yellow), MPS (orange), MPS2 (green), and MPS2+ (blue) (Panel B).

Measures of diagnostic performance for clinically significant prostate cancer were determined for PSA, the PCPTrc, MPS, and the locked MPS2 and MPS2+ models (**Figure 3B**). The AUC of PSA, the PCPTrc, MPS, MPS2, and MPS2+ were 0.597 (95% confidence interval [CI], 0.547 to 0.646), 0.659 (95% CI, 0.611 to 0.707), 0.737 (95% CI, 0.694 to 0.780), 0.807 (95% CI, 0.769 to 0.846), and 0.818 (95% CI, 0.781 to 0.855), respectively. Relative to PSA, the PCPTrc, and MPS, the MPS2+ model improved AUC for clinically significant prostate cancer by 22.1%, 15.9%, and 8.1%, respectively.

While increased AUC demonstrates improved overall diagnostic accuracy (i.e., across all possible threshold values), we sought to evaluate test performance under clinically applicable testing approaches providing high sensitivity for clinically significant cancer. For each test, we identified the threshold value (i.e., cutoff) providing 95% sensitivity, and we calculated the associated specificity, NPV, and PPV. Notably, a testing approach with 95% sensitivity results in a 5% rate of missed or delayed diagnoses relative to performing biopsy in all men, and the associated specificity equals the proportion of unnecessary biopsies (those negative for cancer or overdiagnosing grade group 1 cancer) avoided under the testing approach. At the testing threshold providing 95% sensitivity for clinically significant cancer, the specificity of PSA, the PCPTrc, MPS, MPS2, and MPS2+ were 11%, 20%, 23%, 37%, and 41%, respectively (**Table 2**). Notably, MPS2 provided 97% NPV for clinically significant prostate cancer.

**Table 2.**
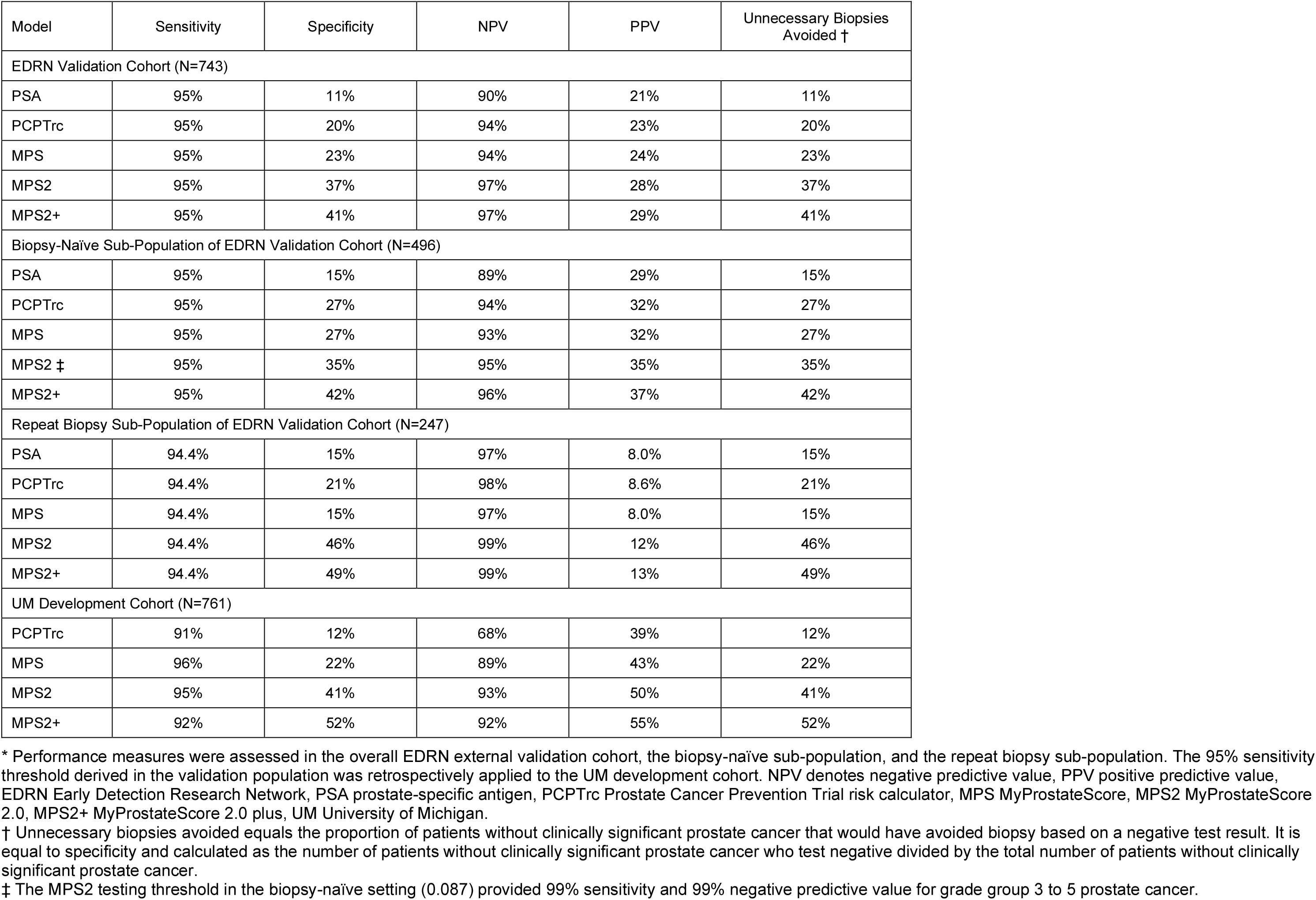
Clinical performance measures of serum PSA, the PCPT risk calculator, MyProstateScore, and MyProstateScore 2.0 models at testing thresholds providing 95% sensitivity for clinically significant prostate cancer.*

### CLINICAL TESTING PARADIGM

Patients with a previous negative prostate biopsy harbor a reduced risk of clinically significant cancer relative to biopsy-naïve patients,^41–43^ meriting inclusion of previous biopsy status at the time of clinical evaluation.^32^ In the proposed diagnostic pathway, under which biomarker testing is performed prior to mpMRI or biopsy, prostate volume (which requires mpMRI or TRUS) is unknown for most biopsy-naïve patients undergoing evaluation. By contrast, prostate volume is available for most patients who have undergone a previous biopsy. In light of this, we evaluated clinical testing strategies based on use of the MPS2 test (without prostate volume) in biopsy-naïve patients and the MPS2+ test (with prostate volume) in those with a prior negative biopsy.

In the biopsy-naïve validation population (N=496), a MPS2 testing approach associated with 95% sensitivity for clinically significant cancer provided 95% NPV and 35% specificity. As compared to PSA, the PCPTrc, and MPS, clinical use of MPS2 would have resulted in absolute reductions of unnecessary biopsies by 20%, 8.0%, and 8.0%, respectively (**Table 2**). While standard clinical data are routinely available in the majority of patients, we found that exclusion of these factors (i.e., using a MPS2 biomarkers-only model) also provided 95% NPV and 35% specificity (**Supplementary Appendix Table S7**). Thus, the absence of one or more components of the clinical history does not appear to reduce the accuracy of MPS2 testing. Notably, MPS2 testing provided 99% sensitivity and 99% NPV for grade group 3 or higher cancer. Full performance measures of MPS2 models in the biopsy-naïve population across clinically pertinent thresholds are included in **Supplementary Appendix Table S7.**

In the repeat biopsy population (N=247), the MPS2+ threshold associated with 95% sensitivity provided 99% NPV and 49% specificity (**Table 2**). Accordingly, use of MPS2+ would have avoided nearly one half of unnecessary biopsies performed under the traditional diagnostic approach. It is notable that the MPS2 model not including prostate volume performed similarly to MPS2+, such that failure to obtain a previous measure of prostate volume does not appear to compromise test performance. Full performance measures of MPS2 models across pertinent threshold values in the repeat biopsy population are included in **Supplementary Appendix Table S8**. Notably, applying the 95% sensitivity thresholds identified in the validation cohort to the development cohort provided similarly high performance (**Table 2**), further supporting robustness of the assay.

### DECISION CURVE ANALYSIS

Decision curve analysis (DCA) was used to evaluate the net clinical benefit of pre-biopsy risk stratification using PSA, the PCPTrc, MPS, and MPS2 models relative to baseline approaches in which all patients undergo biopsy or no patients undergo biopsy. As illustrated in **Supplementary Appendix Figure S6A**, PSA and the PCPTrc provided no net benefit relative to biopsying all patients at risk thresholds less than 15%. While the MPS test provided net clinical benefit across threshold probabilities, the MPS2 and MPS2+ tests provided the highest clinical benefit across all clinically pertinent threshold probabilities. This relationship is further highlighted in **Supplementary Appendix Figure S6B**, which provides the net reduction in biopsies performed based on use of each test. Again, the MPS2 models provided the greatest net reduction in biopsies performed per 100 patients. At the threshold probability of 20%, risk stratification with PSA, the PCPTrc, MPS, MPS2, and MPS2+ would have reduced biopsies performed by 5, 11, 20, 38, and 38, respectively. Reductions in biopsies performed for each test at pertinent threshold probabilities are listed in **Supplementary Appendix Table S9**.

## DISCUSSION

When diagnosed at an early stage, prostate cancer is imminently curable with treatment, and large, randomized clinical trials have demonstrated reduced cancer mortality with population-wide PSA screening.^2–5^ Nonetheless, assessments of PSA screening programs have concluded there is limited evidence of net benefit, secondary to the harms associated with frequent negative biopsies and widespread overdiagnosis of indolent, low-grade cancers.^7–9^ Based on this uncertain benefit-to-harm ratio, trends in practice away from screening have resulted in a rising incidence of incurable, metastatic prostate cancer for the first time in decades.^14, 44^ Despite a clinical capacity to detect and effectively treat prostate cancer that is uniquely high among prevalent malignancies, the lack of a practical, highly accurate, and well-defined diagnostic approach in men with elevated serum PSA has impeded progress against the disease, which remains a leading cause of morbidity and mortality worldwide. Ultimately, there is an extraordinary need for a reliable clinical test to reduce the harms associated with prostate cancer screening and preserve its known mortality benefits.

Translating sequencing-based discovery efforts to an expandable multiplex qPCR platform, we developed and validated a novel urinary test capturing genetic markers specifically associated with prostate cancer, and – for the first time – high-grade prostate cancers at the focus of contemporary clinical practice. Overall, we performed multiplex molecular testing of 1,623 clinical urinary specimens, representing the largest such effort to our knowledge. In an external validation population indicated for biomarker testing, we found that the novel MPS2 test provided substantial diagnostic improvement relative to validated PSA-based tools and the original MPS test, which is one of six biomarker tests currently offered by clinical guidelines.^10^ In patients traditionally referred for prostate biopsy, a MPS2 testing approach providing 95% sensitivity for clinically significant cancer offered 97% NPV and would have avoided between 37% and 41% of unnecessary prostate biopsies. After stratifying patients based on history of previous biopsy, a negative MPS2+ test provided 99% NPV and would have avoided approximately one-half of unnecessary biopsies in men with a previous history of negative biopsy (**Table 2**).

For individual patients, negative predictive values approaching 100% provide clear clinical guidance and unprecedented reliability, allowing for confident, data-driven decision making. For clinicians, reflex use of MPS2 at the practice- or population-level would allow for avoidance of up to one-half of unnecessary biopsies, while preserving immediate detection of 95% of clinically significant cancers diagnosed under the traditional, highly morbid “biopsy all” approach. Critically, MPS2 testing provided 99% sensitivity and NPV for grade group 3 and higher cancers. In other words, the rare cases in which MPS2 testing would have missed or delayed detection of clinically significant cancer (i.e., false negatives) were almost uniformly grade group 2 cancers.^45^ These data provide strong support for use of MPS2 as a practical, highly accurate secondary test in men with elevated PSA to mitigate the potential harms of screening and preserve its long-term benefits.

The ideal diagnostic test has been described as one that is safe, accurate, available, actionable, and providing a favorable benefit-to-harm ratio.^46, 47^ While serum PSA offers favorable practical attributes, the well described harms associated with PSA testing established the need for a complementary test to improve the benefit profile of screening. Multiparametric MRI has been proposed for this role, and high-quality data have shown that pre-biopsy mpMRI improves detection of clinically significant cancer in men with a positive mpMRI.^48, 49^ On the other hand, data describing the use of negative mpMRI to rule out clinically significant cancers range from middling – a pooled 91% NPV (95% confidence interval, 88% to 93%) at the most experienced centers – to highly concerning, with NPV as low as 63% at one expert center and 40% among individual academic radiologists.^11^ Notably, a 2020 systematic review found insufficient published data to even calculate NPV at community hospitals, where the vast majority of the population receives care.^11^ Moreover, the increased cost, time and resource burden, and subjective interpretation of mpMRI present several practical barriers to its population-level use as a first-line test following PSA. While mpMRI is a valuable addition to the diagnostic armamentarium, both practical and performance characteristics suggest it may be best suited later in the diagnostic pathway, e.g., to improve the yield of needle biopsy in men most likely to benefit from invasive testing.

By contrast, biomarker tests provide objective risk quantification and are readily obtained in the standard clinical setting. Building upon previous blood- and urine-based assays including two to four cancer-associated biomarkers, the MPS2 assay uniquely captures data spanning 17 cancer- and high-grade cancer-associated genes. In addition to highly accurate pre-biopsy risk prediction, arming patients and clinicians with early, non-invasive molecular cancer data opens the door to more informed, individualized cancer care across the diagnostic and therapeutic settings. For example, in patients indicated for biopsy after MPS2 testing, the association of tumor subtypes with mpMRI visibility suggests that molecular data could distinguish patients that stand to benefit from pre-biopsy mpMRI from those that should proceed directly to standard biopsy.^50^ In patients with low-grade cancer on biopsy, expression of high-grade genes in pre-biopsy urine would suggest the presence of an occult, aggressive tumor not sampled on diagnostic biopsy, prompting early repeat biopsy.^51^ While prostate biopsy is limited by sampling, urine provides a comprehensive assessment of overall prostatic gene expression – an ideal complement to offset the main limitation of needle biopsy. Subsequently, during surveillance of low-grade cancers, where cancer-specific markers offer limited actionable data, serial assessment of high-grade markers could provide a reliable trigger for biopsy, reducing the need for routine biopsies that remain an undesirable component of active surveillance. Finally, leveraging the association of specific molecular pathways (e.g., DNA damage response) with response to common treatments, including radiotherapy and androgen deprivation therapy,^52, 53^ molecular data from a pre-diagnostic MPS2 test could feasibly inform downstream treatment decisions following detection of aggressive cancer. This level of tumor-specific clinical guidance has not previously been plausible with non-invasive testing. Further characterization of MPS2 genes or the addition of previously validated predictive biomarkers to the MPS2 testing panel are two highly feasible mechanisms for actualizing these strategies in the short-term.

The current study has notable limitations. For one, there was limited racial diversity in our study population, and molecular subtype expression is known to vary by race.^54^ Thus, it is unclear how our findings could differ in African American men, and we are currently pursuing such analyses to ensure an optimal testing approach for all patients. Second, the current analysis was based on systematic prostate biopsy pathology, which is subject to undersampling and could result in misclassification.^55–57^ For this reason, we repeated our analysis in patients with more definitive pathology (e.g., radical prostatectomy), and there was substantial overlap between models (**Table S4**). Third, the current study does not aim to define the combined use of MPS2 and mpMRI, which could be useful at sites of high mpMRI uptake. However, we are currently conducting a prospective, multi-center trial designed to provide unbiased assessment of these tools in combination.^58^ That notwithstanding, performance of MPS2 in the current analysis supports its use as an initial, standalone test in men with an elevated PSA – to rule out the need for mpMRI and biopsy altogether. Thus, evaluation of MPS2 in combination with mpMRI does not preclude the significance and high potential for clinical impact presented in the current report. Additional studies are underway to corroborate these findings and confirm the positive impact of MPS2 testing on longer-term health outcomes.

## Data Availability

All data produced in the present work are contained in the manuscript.

## ACKNOWLEDGEMENTS

We thank the EDRN-PCA3 Study Group, especially Drs. Martin G. Sanda, Ziding Feng, David H. Howard, Scott A. Tomlins, Lori J. Sokoll, Daniel W. Chan, Meredith M. Regan, Jack Groskopf, Jonathan Chipman, Dattatraya H. Patil, Douglas S. Scher, Jacob Kagan, Sudhir Srivastava, Ian M. Thompson Jr, Jing Fan, Aron Y. Joon, Leonidas E. Bantis, and Mark A. Rubin, for their guidance and efforts in developing the validation cohort used in this study. We thank Dr. Stephanie Ellison for her help in the editing and preparation of this study. This work was funded by the Michigan-Vanderbilt EDRN Biomarker Characterization Center (U2C CA271854), the EDRN DMCC (U24 CA086368), Michigan Prostate SPORE (P50 CA186786), NCI Outstanding Investigator Award (A.M.C., R35 CA231996), Prostate Cancer Foundation Young Investigator Award (J.T., L.X., 20YOUN11), Prostate Cancer Foundation, Howard Hughes Medical Institute, and the American Cancer Society.

## AUTHOR CONTRIBUTION STATEMENT

J.J.T., Y. Zheng, J.T.W., and A.M.C. supervised this study and its related analyses. J.J.T., Y. Zhang, L.X., N.L.S., Y.S.N., and A.M.C. wrote the manuscript. J.J.T., N.L.S., Z.C., J.S., G.H., N.V., and K.A. assembled and curated the UM clinical cohort. Y.S.N. carried out RNAseq analysis and transcript nomination. Y. Zhang and B.J.T. developed the MPS2 models. L.X. and H.Z. processed biospecimens and ran OpenArray analyses. Y. Zheng and C.X. carried out validation analyses on the EDRN cohort. T.M.M., G.S.P., S.S.S., J.S., J.T.W., L.P.K., and A.M.C. supported the UM Urology urine cohort. B.J.T. and Y. Zheng provided biostatistics oversight. T.M.M., G.S.P., S.S.S., and A.E.R. provided clinical input on the study and its design.

