## Supplementary Appendix for "Development and Validation of MyProstateScore 2.0 to Detect Clinically Significant Prostate Cancer"

This appendix has been provided by the authors to give readers additional information about their work.

**Supplementary Appendix**

**Table S1. Characterization of 58,724 Gene Targets Assessed by RNAseq.**

See Excel file, “Table S1_Descriptive data of RNAseq targets”.

**Table S2. RNAseq-Based Marker Nomination Criteria.***

| Criterion | Cancer Criteria | High-Grade Criteria | Alternative High-Grade Criteria |
|---|---|---|---|
| 1 | Gene length >500 bp | Gene length >500 bp | Gene length >500 bp |
| 2 | Differential expression (cancer versus normal) adjusted p-value <0.1 | Differential expression adjusted p-value <0.1 | Differential expression (low-grade versus high-grade cancers) adjusted p-value <0.1 |
| 3 | log2 mean fold-change >0 | log2 mean fold-change >0 | log2 mean fold-change >0.5 |
| 4 | >50% of cancer samples in the high-expressing samples | >85% of high-expressing samples in high-grade cancers via clustering statistics | >80% of high-expressing samples in high-grade cancers via clustering statistics |
| 5 | 95th percentile normal expression <50 TPM | log2 fold-change (high-expressing sample mean versus low-expressing sample mean) >0 | log2 mean expression of high-expressing samples >6 |
| 6 | log2 fold-change ($50^{th}$ percentile cancer sample versus $75^{th}$ percentile normal samples) >0 | $95^{th}$ percentile normal expression <50 TPM | |
| 7 | log2 fold-change ($90^{th}$ percentile cancer sample versus $90^{th}$ percentile normal samples) >1.9 or log2 fold-change ($90^{th}$ percentile cancer sample versus $50^{th}$ percentile normal samples) >3.75 | $95^{th}$ percentile high-grade expression >30 TPM | |

*RNAseq-based analysis for identification of novel informative biomarkers utilized one set of criteria for cancer-associated genes and two sets of criteria for high-grade cancer-associated genes. bp denotes base pairs, log2 log base 2, TPM transcripts per million.

**Table S3. Indications for Prostate Biopsy in the External Validation Cohort.**

| Indication for biopsy ‐ no. (%) | Biopsy-Naïve (N=496) | Repeat Biopsy (N=247) | Total (N=743) |
|---|---|---|---|
| PSA >2.0 ng/ml | 294 (59%) | 141 (57%) | 435 (59%) |
| Elevated PSA Velocity | 95 (19%) | 48 (19%) | 143 (19%) |
| Abnormal DRE | 94 (19%) | 29 (12%) | 123 (17%) |
| Prior ASAP or HGPIN | 0 (0%) | 19 (7.7%) | 19 (2.6%) |
| Percent Free PSA <15% | 9 (1.8%) | 3 (1.2%) | 12 (1.6%) |
| Other | 4 (0.8%) | 7 (2.8%) | 11 (1.5%) |

DRE denotes digital rectal exam, PSA prostate-specific antigen, ASAP atypical small acinar proliferation, HGPIN high-grade prostatic intraepithelial neoplasia.

**Table S4. Model Coefficients from MPS2 Re-Development Using Maximum Grade Group from All Biopsy and Surgical Specimens.***

| Covariate | MPS2 † | RP-Derived MPS2 | MPS2+ ‡ | RP-Derived MPS2+ |
|---|---|---|---|---|
| (Intercept) | 5.902430658 | 5.36963703 | 6.676363594 | 5.66713268 |
| T2ERG | 0.111906862 | 0.12028164 | 0.148584627 | 0.15333868 |
| SCHLAP1 | 0.17335791 | 0.15765111 | 0.205268829 | 0.14709972 |
| OR51E2 | 0.200676934 | 0.17998234 | 0.228791882 | 0.15904006 |
| APOC1 | -0.07916931 | -0.0798262 | -0.08896388 | -0.0924547 |
| PCAT14 | 0.14420976 | 0.05653337 | 0.16009867 | 0.08193915 |
| CAMKK2 | -0.26364401 | -0.2662075 | -0.277941927 | -0.2934813 |
| PCA3.1 | 0.080881661 | 0.12143166 | 0.074209893 | 0.09433227 |
| NKAIN1 | -0.06946207 | N/A | -0.093791082 | N/A |
| B3GNT6 | 0.047475092 | 0.04017777 | 0.072524885 | 0.05564982 |
| TFF3 | 0.186395669 | 0.20852164 | 0.2128103 | 0.24750486 |
| SPON2 | 0.156664808 | 0.08542159 | 0.1740959 | 0.03934708 |
| PCGEM1 | -0.16940833 | -0.1154365 | -0.149084289 | -0.1369629 |
| TRGV9 | 0.096184103 | 0.2130775 | 0.177972309 | 0.24262976 |
| TMSB15A | 0.151071453 | N/A | 0.214870771 | N/A |
| ERG | 0.023544761 | N/A | 0.030085251 | N/A |
| KLK4 | 0.149451849 | N/A | 0.214609188 | N/A |
| HOXC6 | 0.05612131 | 0.07807035 | 0 | 0.06422307 |
| ACSM1 | N/A | 0 | N/A | -5.73E-05 |
| LRRN1 | N/A | -0.0212629 | N/A | 0 |
| MS4A8 | N/A | 0.00093909 | N/A | 0.02808881 |
| GRIN3A | N/A | 0 | N/A | -0.0040861 |
| Age | 0.000134221 | 0.00168473 | 0.021446485 | 0.0210196 |
| African American | 0.828856591 | 0.47329332 | 1.232493234 | 0.56924532 |
| Family History | 0.148709502 | 0.00459593 | 0.292757369 | 0.0820011 |
| Abnormal DRE | 0.888379309 | 0.68733551 | 1.094432439 | 0.86237867 |
| Previous Biopsy | -0.8505938 | -0.6063101 | -0.61694213 | -0.4762385 |
| PSA | 0.073709982 | 0.03565447 | 0.092554335 | 0.10335382 |
| Prostate Volume | N/A | N/A | -0.024051593 | -0.0255555 |

*Considering potential misclassification of clinically significant prostate cancer due to biopsy undersampling, we evaluated MPS2 models including pathologic data obtained subsequent to study urine collection (e.g., repeated biopsy, radical prostatectomy). Of 761 patients in the development set, 382 (50%) underwent additional biopsy (N=201) and/or radical prostatectomy (N=217), and 71 (11%) were upgraded to clinically significant prostate cancer. The table includes parameters from models derived based on the highest cancer grade detected (i.e., RP-derived models). Of the 17 informative markers in the MPS2 model, 13 were retained in the RP-derived model. The direction of biomarker association with the outcome was unchanged for all markers. The AUC of the cross validated models differed by approximately 1% (0.802 vs. 0.792 for MPS2, and 0.821 vs. 0.822 for MPS2+). MPS2 denotes MyProstateScore2.0, MPS2+ MyProstateScore 2.0 plus, RP radical prostatectomy and/or repeat biopsy.

†MPS2: Calibrated logit = -1.453526 + logit*1.302089

‡ MPS2+: Calibrated logit = -1.41207 + logit*1.077061

**Table S5. Characterization of the 54-Marker MPS2 Candidate Panel.**

See Excel file, “Table S5_ Descriptive data on 54 genes evaluated for inclusion in MPS2 model”.

**Table S6. Description of Informative Biomarkers in the MPS2 Models for Clinically Significant Prostate Cancer.**

| No. | Gene Name | Chromosome | Gene ID | Probe | Frequency | Cumulative Importance * |
|---|---|---|---|---|---|---|
| **1** | TMPRSS2-ERG | 21-21 | ENSG00000184012, ENSG00000157554 | Hs03063375_ft | 40 | 1265 |
| **2** | SCHLAP1 | 2 | ENSG00000281131 | Hs04968419_m1 | 35 | 1582 |
| **3** | OR51E2 | 11 | ENSG00000167332 | Hs04231197_m1 | 33 | 2006 |
| **4** | APOC1 | 19 | ENSG00000130208 | Hs00155790_m1 | 31 | 456 |
| **5** | PCAT14 | 22 | ENSG00000280623 | Hs04941925_m1 | 30 | 841 |
| **6** | CAMKK2 | 12 | ENSG00000110931 | Hs00902176_m1 | 29 | 1604 |
| **7** | PCA3 | 9 | ENSG00000225937 | Hs01371939_g1 | 28 | 1015 |
| **8** | NKAIN1 | 1 | ENSG00000084628 | Hs01563334_m1 | 28 | 456 |
| **9** | B3GNT6 | 11 | ENSG00000198488 | Hs00934529_s1 | 28 | 211 |
| **10** | TFF3 | 21 | ENSG00000160180 | Hs00902278_m1 | 26 | 1329 |
| **11** | SPON2 | 4 | ENSG00000159674 | Hs00202813_m1 | 26 | 1080 |
| **12** | PCGEM1 | 2 | ENSG00000227418 | Hs01369007_m1 | 26 | 725 |
| **13** | TRGV9 | 7 | ENSG00000211695 | Hs00233330_m1 | 24 | 955 |
| **14** | TMSB15A | X | ENSG00000158164 | Hs00762927_s1 | 22 | 548 |
| **15** | ERG | 21 | ENSG00000157554 | Hs01554635_m1 | 21 | 221 |
| **16** | KLK4 | 19 | ENSG00000167749 | Hs00191772_m1 | 20 | 1094 |
| **17** | HOXC6 | 12 | ENSG00000197757 | Hs00171690_m1 | 20 | 354 |

*Cumulative importance indicates the relative weight of marker importance summed across repeat samplings as derived by elastic net modeling.

**Table S7. Clinical Performance of High Sensitivity MPS2 Threshold Values in the Biopsy-Naïve External Validation Population (N=496).***

| **Threshold** | **Sensitivity** | **Specificity** | **NPV** | **PPV** | ***Unnecessary Biopsies Avoided*** |
|---|---|---|---|---|---|
| MPS2+ | | | | | |
| ≤0.075 | 96% | 31% | 96% | 34% | 31% |
| ≤0.087 | 95% | 34% | 95% | 34% | 34% |
| *≤0.10* | *95%* | *38%* | *95%* | *36%* | *38%* |
| ≤0.15 | 90% | 53% | 94% | 41% | 53% |
| **MPS2** | | | | | |
| ≤0.075 | 96% | 31% | 96% | 34% | 31% |
| *≤0.087* | *95%* | *35%* | *95%* | *35%* | *35%* |
| ≤0.10 | 94% | 39% | 95% | 36% | 39% |
| ≤0.15 | 90% | 54% | 93% | 41% | 54% |
| MPS2 Biomarkers Only | | | | | |
| *≤0.075* | *95%* | *35%* | *95%* | *35%* | *35%* |
| ≤0.087 | 95% | 34% | 95% | 34% | 34% |
| ≤0.10 | 90% | 44% | 92% | 37% | 44% |

*The proposed MPS2 model for use in the biopsy-naïve population is listed in bold (i.e., MPS2). The optimal threshold value approximating 95% sensitivity proposed for clinical use is shown in italics. Unnecessary biopsies avoided equals the proportion of patients without clinically significant prostate cancer that would have avoided biopsy with use of the proposed test. It is equal to specificity and calculated as the number of patients without clinically significant prostate cancer who test negative divided by the total number of patients without clinically significant prostate cancer. NPV denotes negative predictive value, PPV positive predictive value, MPS2 MyProstateScore 2.0, MPS2+ MyProstateScore 2.0 plus.

**Table S8. Clinical Performance of High Sensitivity MPS2 Threshold Values in the Repeat Biopsy External Validation Population (N=247).***

| Threshold | Sensitivity | Specificity | NPV | PPV | *Unnecessary Biopsies Avoided* |
|---|---|---|---|---|---|
| **MPS2+** | | | | | |
| *≤0.054* | *94.4%* | *49%* | *99%* | *13%* | *49%* |
| ≤0.065 | 89% | 55% | 98% | 13% | 55% |
| ≤0.075 | 83% | 57% | 98% | 13% | 57% |
| ≤0.10 | 67% | 62% | 96% | 12% | 62% |
| MPS2 | | | | | |
| *<0.045* | *94.4%* | *46%* | *99%* | *12%* | *46%* |
| <0.06 | 89% | 52% | 98% | 13% | 52% |
| <0.065 | 78% | 55% | 97% | 12% | 55% |
| ≤0.10 | 67% | 68% | 96% | 14% | 68% |
| MPS2 Biomarkers Only | | | | | |
| ≤0.07 | 100% | 35% | 100% | 11% | 35% |
| *≤0.08* | *94.4%* | *41%* | *99%* | *11%* | *41%* |
| ≤0.10 | 78% | 48% | 97% | 11% | 48% |

*The proposed MPS2 model for use in the repeat biopsy population is listed in bold (i.e., MPS2+). The optimal threshold value approximating 95% sensitivity proposed for clinical use is shown in italics. Unnecessary biopsies avoided equals the proportion of patients without clinically significant prostate cancer that would have avoided biopsy with use of the proposed test. NPV denotes negative predictive value, PPV positive predictive value, PSA prostate-specific antigen, PCPTrc Prostate Cancer Prevention Trial risk calculator, MPS MyProstateScore, MPS2 MyProstateScore 2.0, MPS2+ MyProstateScore 2.0 plus.

**Table S9. Decision Curve Analysis – Net Reduction in Biopsies Performed per 100 Patients.***

| | Threshold Probability | | | |
|---|---|---|---|---|
| | **5%** | **10%** | **15%** | **20%** |
| **PSA** | 0 | 0 | 0 | 5 |
| **PCPTrc** | 0 | 0 | 0 | 11 |
| **MPS** | 1 | 7 | 14 | 20 |
| **MPS2** | 7 | 23 | 32 | 38 |
| **MPS2+** | 12 | 22 | 31 | 38 |

*Reduction in biopsies performed at pertinent threshold probabilities without missing a single diagnosis of clinically significant prostate cancer. The threshold probability reflects how the patient and clinician value potential clinical outcomes. For example, a threshold probability of 5% applies to patients that would choose to pursue biopsy if their risk of clinically significant cancer is 5% or higher. This implies a highly risk-averse population, such as younger men with a long life-expectancy. A threshold probability of 20% applies to patients that would choose to pursue biopsy only if their risk of clinically significant cancer is 20% or higher. Such a population strongly values avoiding biopsy and is willing to accept a higher risk of delayed detection of clinically significant cancer. An optimal marker provides the highest benefit across a range of reasonable preferences. PSA denotes prostate-specific antigen, PCPTrc Prostate Cancer Prevention Trial risk calculator, MPS MyProstateScore, MPS2 MyProstateScore 2.0, MPS2+ MyProstateScore 2.0 plus.

**Figure S1. Heatmap Illustrating Differential Expression of RNAseq Target Genes.**

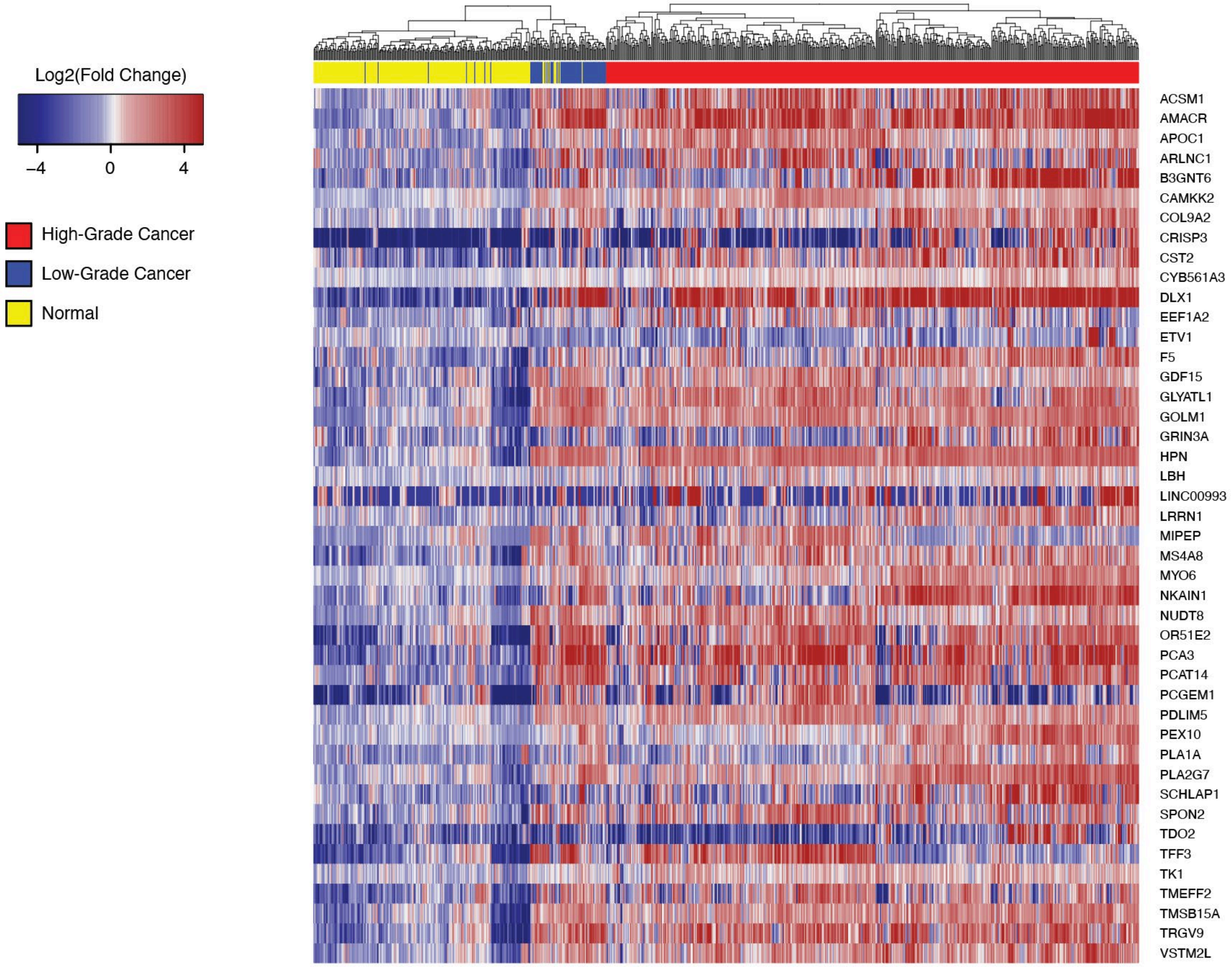

Legend: Shown is a heatmap illustrating differential expression of 44 target genes in the discovery set as determined by RNAseq. The color bar below the column-wise hierarchical clustering tree represents sample pathology. Normal samples, defined as those with no cancer, are shown in yellow (N=220), low-grade cancers are shown in blue (N=71), and higher-grade cancers (grade group 2 or higher) are shown in red (N=484). Red, white, and blue cells in rows represent differential gene expression, calculated as $\log_2$(fold change in gene expression) divided by mean expression in benign samples. Dark blue represents decreased expression relative to normal, red represents increased expression relative to normal, and white represents no difference. The column-wise dendrogram represents hierarchical clustering among high-grade and low-grade cancer to normal samples.

**Figure S2. Box and Dot Plots of Gene Expression in Benign, Low-Grade Cancer, and High-Grade Cancer Tissue in the Discovery Set.**

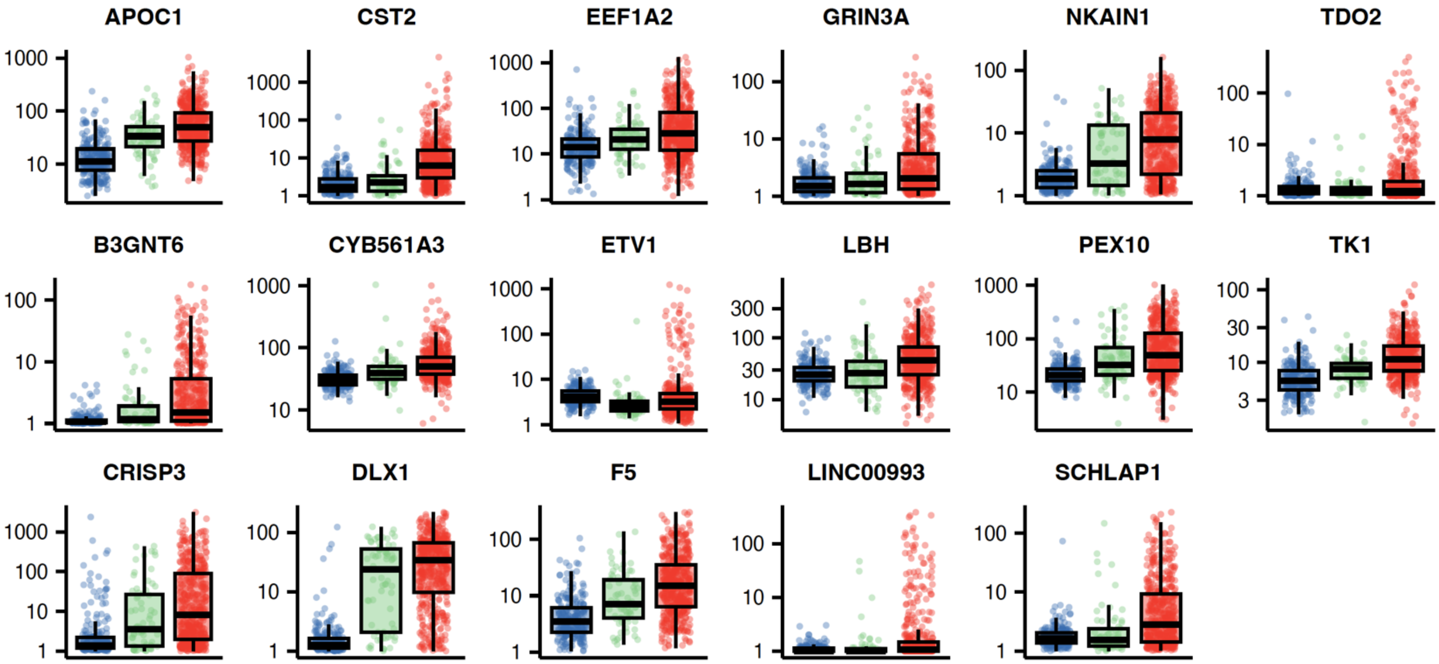

Legend: Shown are box and dot plots of gene expression from tissue-based RNAseq analyses for the 17 biomarkers meeting pre-defined nomination criteria for high-grade prostate cancer. Plots illustrate log-transformed RNAseq-derived transcripts per million (TPM) for benign prostate tissue (blue), low-grade prostate cancer (green), and higher-grade prostate cancer (red).

**Figure S3. Box and Dot Plots of Gene Expression in Benign Versus Prostate Cancer Tissue in the Discovery Set.**

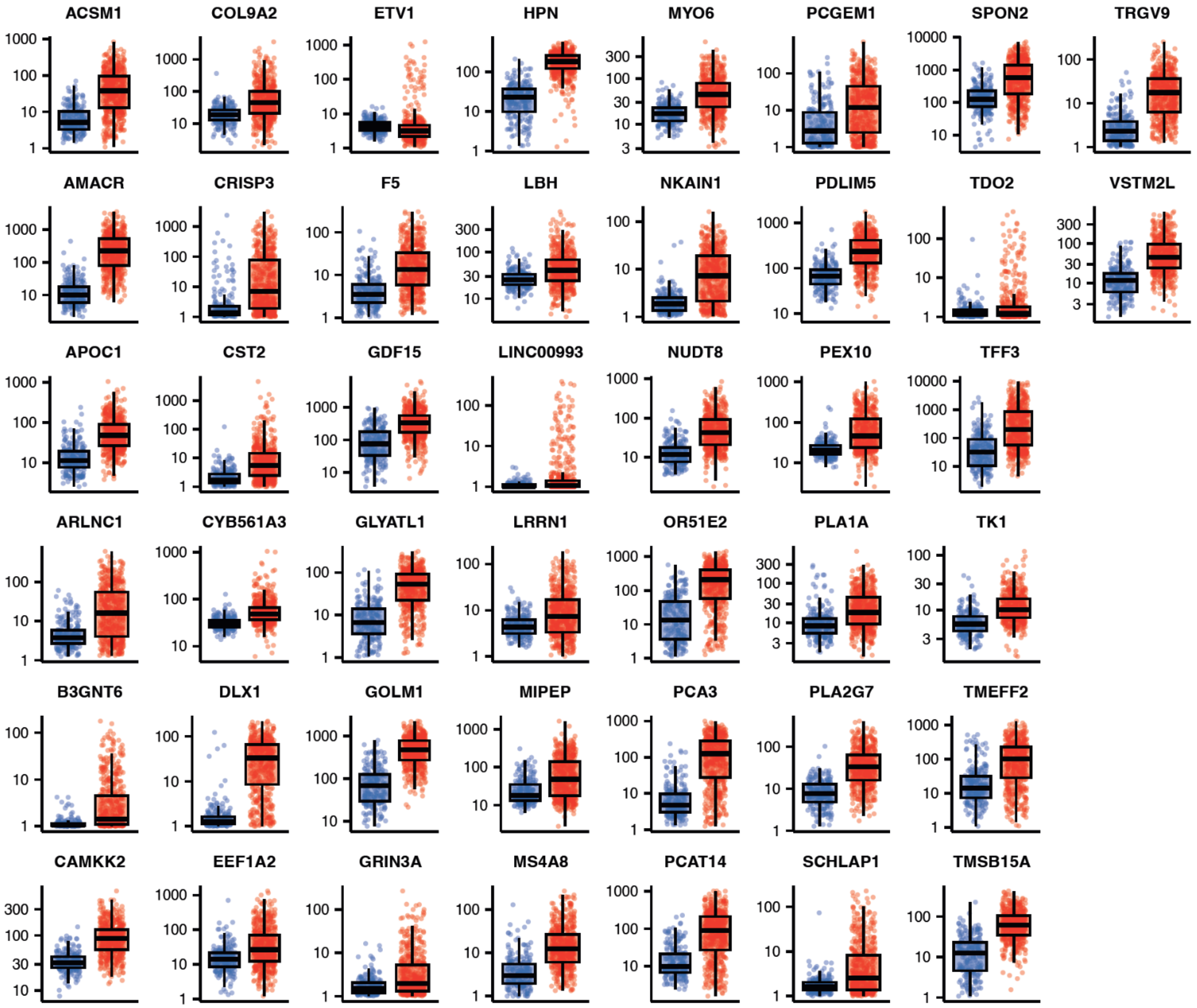

Legend: Shown are box and dot plots of gene expression from tissue-based RNAseq analyses for the 44 biomarkers meeting pre-defined nomination criteria for prostate cancer. Plots illustrate log-transformed RNAseq-derived transcripts per million (TPM) for benign prostate tissue (blue) and prostate cancer (red).

**Figure S4. Flow Diagram of the University of Michigan Development Cohort.**

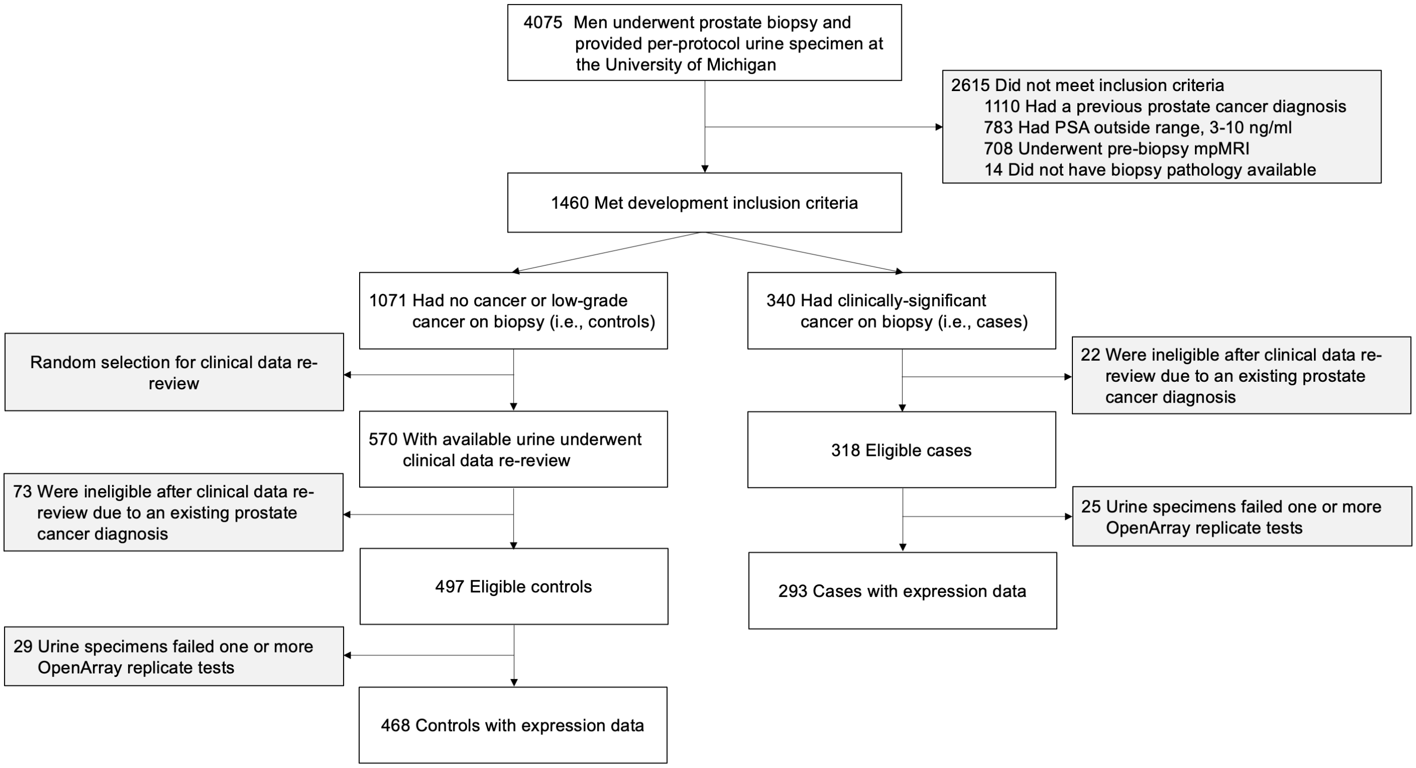

Legend: Among 1460 eligible subjects in the University of Michigan biospecimen database, 340 were found to have clinically significant cancer on biopsy (i.e., cases). Power analysis demonstrated a 1:1 case to control (i.e., no cancer or grade group 1 cancer on biopsy) design with N=300 per group provided >95% power to detect differential expression of up to 20 candidate biomarkers. Considering the incremental power gained from additional controls, we assessed 570 eligible controls with adequate urine for study inclusion. After re-review of clinical data, 73 controls (13%) were excluded due to history of a positive biopsy prior to specimen collection. The OpenArray™ assay failed one or more replicates in 29 eligible controls and 25 eligible cases, yielding the final cohort for analysis. PSA denotes prostate-specific antigen, mpMRI multi-parametric magnetic resonance imaging, MPS2 MyProstateScore 2.0.

**Figure S5. Pre- and Post-Calibration Curves of MPS2 and MPS2+ in the Development Cohort.**

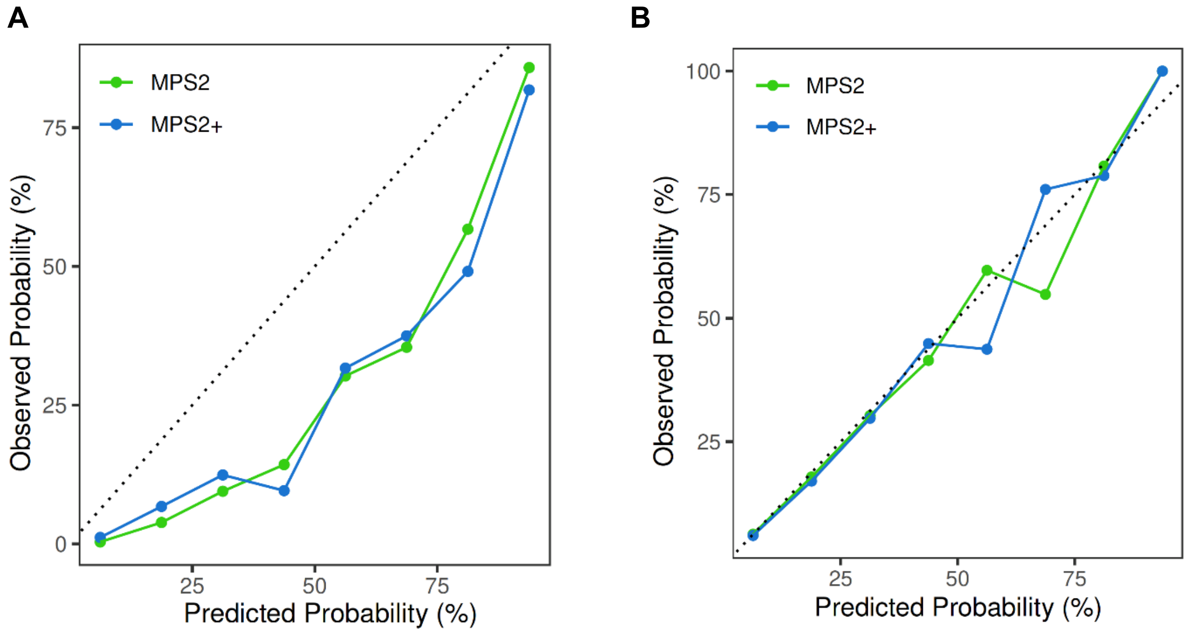

Legend: Shown are the pre-calibration (Panel A) and post-calibration (Panel B) curves for MPS2 (green) and MPS2+ (blue) in the development cohort after re-sampling to match clinically significant cancer prevalence in the validation cohort (i.e., 20%). Calibration involved re-estimation of model intercept and slope and was performed using the *calibration* function from the R package *caret*.

**Figure S6. Decision Curve Analysis Assessing (A) Net Benefit and (B) Net Reduction in Biopsies using MPS2 and Other Testing Strategies.**

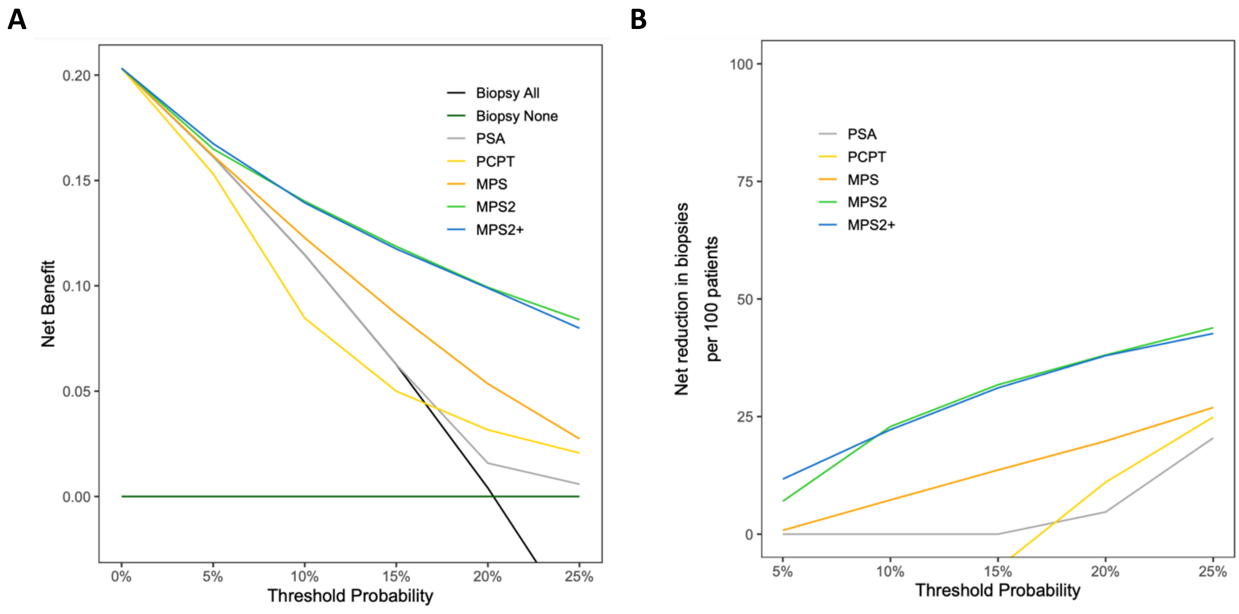

Legend: **A.** Shown are decision curve analysis (DCA) plots for net clinical benefit of pre-biopsy testing with PSA (gray), the PCPTrc (yellow), MPS (orange), MPS2 (green), and MPS2+ (blue) as compared to baseline approaches of “biopsy all” (black) or “biopsy none” (dark green). The threshold probability (x-axis) reflects how the patient and clinician value potential clinical outcomes. For example, a threshold probability of 5% applies to patients that would choose to pursue biopsy if their risk of clinically significant cancer is 5% or higher. At a practice level, this implies that the clinician would be willing to perform as many as 20 biopsies to detect an additional clinically significant cancer. For clinically significant prostate cancer, a 5% threshold probability represents a highly risk-averse population, such as younger men with a long life-expectancy. At the other end of the spectrum, a threshold probability of 20% applies to patients that would choose to pursue biopsy only if their risk of clinically significant cancer is ≥20%. Such a population more strongly values avoiding biopsy and is willing to accept a higher risk of delayed detection of clinically significant cancer. As illustrated in the figure, the MPS2 and MPS2+ models provide the highest net benefit across the entire range of clinically pertinent threshold probabilities. **B**. Shown are DCA plots illustrating the net reduction in biopsies performed per 100 patients (without missing a single diagnosis of clinically significant prostate cancer) based on pre-biopsy testing using PSA (gray), the PCPTrc (yellow), MPS (orange), MPS2 (green), and MPS2+ (blue) as compared to a baseline approach of biopsying all at-risk patients. The MPS2 and MPS2+ models provide the largest reduction in biopsies performed across all clinically pertinent threshold probabilities.
